# Prevalence of adequately iodized salt and its determinants in Gambela district, Southwest Ethiopia

**DOI:** 10.1101/2023.01.17.23284676

**Authors:** Getamesay Aynalem Tesfaye, Ebissa Negera Gemechu, Abdella Yasin Umer, Fentaneh Teshome Chanie

## Abstract

**Background:** Iodine deficiency disorders (IDDs) are among the major public health problems for populations all over the world. Even though the recommended strategy for IDD control is universal salt iodization, little was known about availability of adequately iodized salt in Gambela region.

**Objective:** This study was conducted to determine availability of adequately iodized salt and associated factors in Gambela district, Southwest Ethiopia.

**Methods:** A community-based cross-sectional study was conducted in July 2019 in Gambela district. Multistage sampling method was employed to select 422 households. A structured questionnaire and rapid test kits were data collection instruments. Multivariate logistic regression was used to identify association.

**Results:** About 52.8% (95% CI: 0.47, 0.57) of households had adequately iodized salt. College diploma and above educational status (AOR=4.27, 95% CI: 1.47, 12.44), favorable attitude (AOR=5.69, 95% CI: 2.83, 11.42), walking less than 30 minutes to get iodized salt (AOR=4.69, 95% CI: 2.56, 8.60), using packed salt (AOR=2.75, 95% CI: 1.54, 4.93) and using covered salt container (AOR=3.93, 95% CI: 2.21, 6.98) were factors positively associated with availability of adequately iodized salt.

**Conclusions:** The availability of adequately iodized salt in Gambela district is very low. Enhancing community awareness about the benefit and good practice of iodized salt should be emphasized besides improving accessibility.

## Introduction

World Health Organization (WHO) stated that iodine deficiency disorders (IDDs) are among the major public health problems for populations all over the world, as they are a threat to the social and economic development of countries.^1^ Iodine deficiency affects all populations, however, pregnant women, lactating women, women of reproductive age, and children younger than three years of age are considered the most important groups in which to diagnose and treat iodine deficiency.^2^

IDDs during fetal and neonatal growth and development leads to irreversible damage of the brain and central nervous system and, consequently, to irreversible mental retardation.^2^ The most devastating effect of iodine deficiency are increased perinatal mortality and mental retardation.^1^ About 38 million newborns in developing countries every year remain unprotected from the lifelong consequences of brain damage associated with iodine deficiency disorders, which affects a child’s ability to learn, and later in life, to earn; therefore, preventing children, communities and nations from fulfilling their potential.^3^

In Ethiopia, one out of every 1000 is a cretin mentally handicapped, due to congenital iodine deficiency, and about 50 000 prenatal deaths are occurring annually due to IDD.^4^ Almost 35% of children in Ethiopia suffered from goiter, with a highest prevalence of 71% in the South Nations Nationalities and Peoples region.^5^ The prevalence of palpable and visible goiter among pregnant and lactating mothers in Ethiopia was 30.9%.^6^

A diet low in iodine is the main cause of IDD which usually occurs among populations living in areas where the soil has been depleted of iodine because of flooding, heavy rainfall or glaciation.^3^ The recommended strategy for IDD control is based on correcting the deficiency by increasing iodine intake through supplementation or food fortification, especially universal salt iodization (USI) which is the iodization of salt for both human and livestock consumption.^1^ In 1994, WHO and United Nations Children’s Fund (UNICEF) Joint Committee on Health Policy recommended USI as a safe, cost-effective, and sustainable strategy to ensure sufficient intake of iodine by all individual.^7^

Globally, only 75% of the households use iodized salt which had a dramatic increment compared to the 1990 report, 10%.^8^ In sub-Saharan Africa, 64% of households are using iodized salt, nevertheless the level of utilization widely varies from 10 to 90% in different countries.^9^

Ethiopia passed a comprehensive salt regulation in 2011 which mandated that all salt for human consumption in the country should be iodized.^10^ Despite these attempts, iodized salt coverage in Ethiopia was still below the WHO USI target of 90% coverage.^7,11^ Different studies in Ethiopia showed that the availability of adequately iodized salt at household level was much lower than the international USI target.^12-14^

Worldwide various studies revealed different factors associated with availability of adequately iodized salt. Hence, household coverage of adequately iodized salt was significantly higher in households with a higher socioeconomic status than households with a lower socioeconomic status in Bangladesh, Ghana, India, Indonesia, the Philippines, Senegal and Tanzania.^15^ A study in Nepal showed that mean and median iodine concentration of powder (packed) salt was higher than coarse (unpacked) salt.^16^ Elsewhere a study in Pakistan, illiteracy, no knowledge of iodized salt, non-availability of iodized salt, unawareness about the advantages of use of iodized salt were factors associated with non-use of iodized salt for cooking at household.^17^ Community size, educational level, marital status, salt type, access to information, and wealth status were determinants of availability of iodized salt in households in Ghana.^18-19^ Different studies in Ethiopia reported that using packed salt, access to information, income of households, education, knowledge about iodized salt, not exposing salt to sunlight, storing salt in dry place and storing salt in container with a lid were among factors significantly associated with availability of adequately iodized salt.^20-22^

There was Ethiopian national survey conducted in Gambela region on household iodized salt coverage, but the it did not determine factors associated with the availability of adequately iodized salt in the district.^11^ Furthermore, studies conducted on availability of adequately iodized salt in Ethiopia had inconsistent findings.^23-25^ Therefore, this study aimed at determining prevalence of adequately iodized salt and its determinants in Gambela district. Eventually, this study will be big input to health professionals and policy makers for planning and evaluation of the availability of adequately iodized salt at household level for prevention of IDDs.

## Materials and Methods

### Study design

A community-based cross-sectional study was conducted from 1^st^ July 2019 to 31^st^ July 2019.

### Study setting

The study was conducted in Gambela district which is the capital district of Gambela region, one of the nine ethnic divisions of Ethiopia, located 777 kilo meter in the southwest of Addis Ababa.^26^ The district is characterized by hot and humid climate. In the district, there was one government hospital, one government health centre, five private lower clinics, and twelve private medium clinics that were providing health and medical services for the community. In 2019, the district had a total population of 59,462 comprised of 12,927 households, classified in to five kebeles which are smallest administrative unit in Ethiopia.

### Study population

The source population was all households in Gambela district, while the study population was all selected households found in selected kebeles of the district. A member of a household who is 18 years old and above, mostly involved in cooking food, were interviewed. An individual in a selected household who is sick and unable to respond at the time of data collection were excluded from the study.

### Sample size determination

Sample size was calculated by the following assumptions: two-sided confidence level of 95%, power of 80%, exposed to unexposed ratio 1:1, using Epi Info-7 Stat Calc computer software for double population proportions formula, considering 10% for non-response, the design effect of two; and taking from study done in southwest Ethiopia magnitude of availability of adequately iodized salt at household level among those who place salt at a dry place of 30.40% and 2.13 odds ratio.^27^ Finally, the sample size calculated for this study was 422 households.

### Sampling procedure

A multistage sampling method was employed to select households. Accordingly, first 3 kebeles were selected using simple random sampling technique. Then, the sample was allocated proportional to the household size of each administrative area. The first household from each administrative area was identified using lottery method, and then, systematic random sampling technique was applied to identify the next household to be studied.

### Data collection

The data was collected by using structured validated interviewer-administered questionnaire and rapid iodine salt testing. The questionnaire, which was adapted from available literatures, was used for collecting data regarding the household socio-demographic characteristics, knowledge and attitude regarding adequacy of iodized salt and various practices related to the use of iodized salt.^12,22,27^ The adapted questionnaire was modified and contextualized to fit the local situation and the research objective. The questionnaire was initially prepared in English and then translated in to Amharic language then back to English by fluent speakers to check its consistency.

The iodine content of salt in a household was determined by using the rapid iodine test kit, at the end of the interview. Determination of iodine content of iodized salt done by using standardized procedures recommended by the WHO. First of all, one drop of starch solution was squeezed onto a half-teaspoon sample of table salt obtained in each household. If the color changed (from light blue to dark violet), it was matched to a color chart provided with the test kit and the iodine concentration classified as < 15 or ≥ 15 parts per million (PPM). If the initial test was negative (no change in color), a second confirmatory test, adding an acid-based solution in addition to the starch solution, was done. If the color of the salt did not change even after the confirmatory test, the salt sample was considered to contain no iodine.^7^

A household declared to have adequately available iodized salt when the its sampled salt has iodine ≥ 15 PPM.^28^ When the participant answers correctly more than 50% of knowledge questions, the participant has good knowledge.^29^ Favorable attitude and good practice were defined as answering correctly more than 50% of questions about attitude and practice, respectively.^30^

### Data quality control

Before data collection, pre-testing was carried out on 5% of households in the kebeles not selected for the study. Consequently, necessary modifications and corrections were made on the questionnaire before it was used for the actual study. The validity of rapid test kit was routinely monitored. The expiry date of the rapid test kit was checked before use. Training was given to research assistants on the topic and purpose of the research, on how to approach study subjects and how to use the questionnaire and the rapid test kit. The collected data was checked for the completeness, accuracy and clarity by the principal investigator and supervisor. This quality checking was done on the data collection day and after data collection. Correction was made before the next data collection. Data clean-up and cross-checking was done before analysis. Multivariate logistic regression analysis was used to control possible confounding variables. The fitness of multivariate logistic regression was assessed by Hosmer-Lemeshow goodness of fit test.

### Data analysis

The collected data were explored for missing data, and distribution of outcome variable as well as test of parallel lines and model fitness information were checked. Data entry and analysis was done by Statistical Package for the Social Sciences version 20 (SPSS v20) after data cleaning. Descriptive analysis results were presented using tables and texts. Since the interest was identifying households with adequately iodized salt, the dependent variable was coded as one if household sampled salt has iodine ≥ 15 PPM, and coded as zero if not. Multi-collinearity effect was checked and variables with standard error above two were removed from analysis. Those variables with no collinear effect were included in binary logistic regression model, with backward stepwise likelihood-ratio method, to see the possible association with availability of adequately iodized salt. Then, variables with p-value <0.25 in the bivariate analysis were taken as candidate for multivariate analysis. Multivariate logistic regression analysis, with backward stepwise likelihood-ratio method, was employed to identify the predictors of availability of adequately iodized salt. The multivariate logistic regression model was fit according to Hosmer-Lemeshow goodness of fit test (p-value= 0.11). Crude odds ratio and adjusted odds ratio along with their respective 95% Confidence interval was calculated to measure the strength of the association. Association between the explanatory and the dependent variable was assessed at a p-value of 0.05.

### Ethical considerations

An application for full ethical approval was made to the Mettu University Human Research Ethical Review Committee and ethical consent was received on 25 June 2019. The ethics approval number is MUPHD/143/2019. Confidentiality was assured and participants signed on informed consent form.

## Results

### Socio-demographic characteristics

A total of 411 households were interviewed, yielding a response rate of 97.4%. The mean (±standard deviation) age of the participants was 28.9 (±9.7) years. Majority (76.9%) of study participants were female and more than half (61.1%) of respondents’ relationship in the household were mother. A hundred and forty-eight (36.0%) of participants had primary education, whereas 81 (19.7%) of their partner had secondary education. Majority (74.7%) of the study participants lived in male-headed households, and four fifth (81.0%) of study participants had family size less than six (Table 1).

**Table 1:**
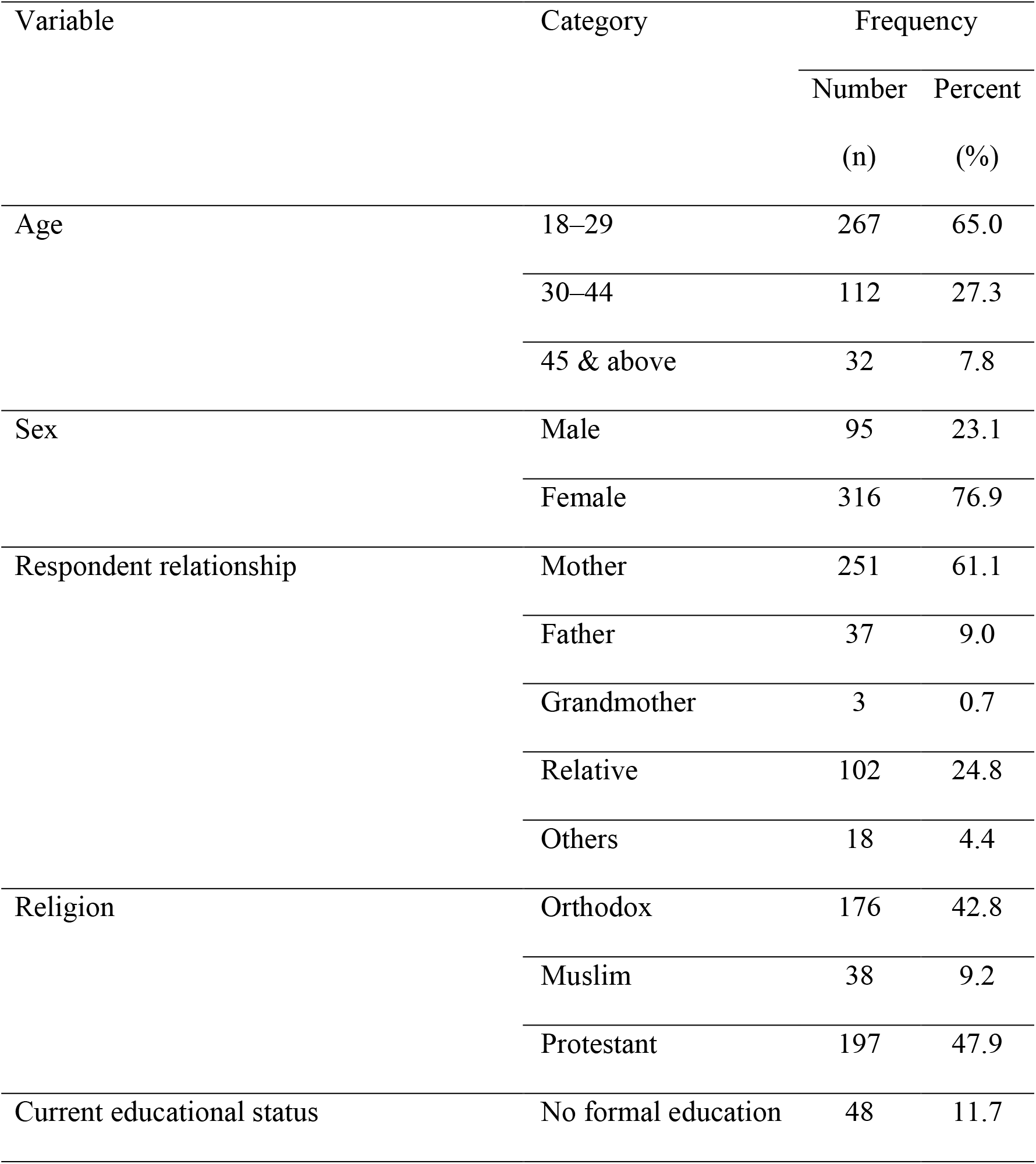
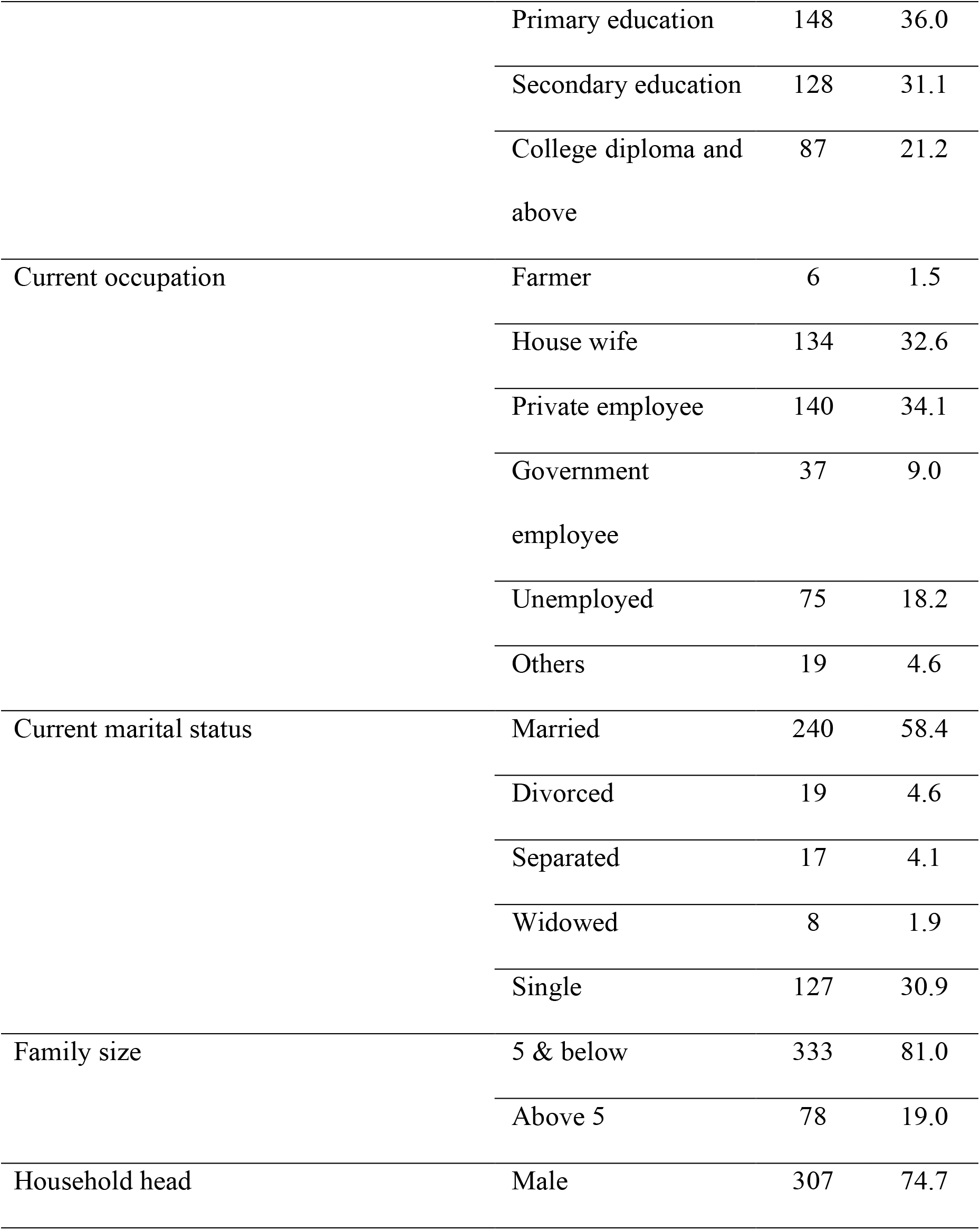
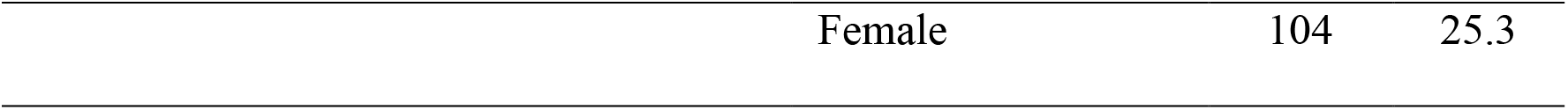
Socio-demographic characteristics of respondents in Gambela district (N= 411).

| Variable | Category | Frequency |  |
| --- | --- | --- | --- |
|  |  | Number | Percent |
|  |  | (n) | (%) |
| Age | 18–29 | 267 | 65.0 |
|  | 30–44 | 112 | 27.3 |
|  | 45 & above | 32 | 7.8 |
| Sex | Male | 95 | 23.1 |
|  | Female | 316 | 76.9 |
| Respondent relationship | Mother | 251 | 61.1 |
|  | Father | 37 | 9.0 |
|  | Grandmother | 3 | 0.7 |
|  | Relative | 102 | 24.8 |
|  | Others | 18 | 4.4 |
| Religion | Orthodox | 176 | 42.8 |
|  | Muslim | 38 | 9.2 |
|  | Protestant | 197 | 47.9 |
| Current educational status | No formal education | 48 | 11.7 |
|  | Primary education | 148 | 36.0 |
|  | Secondary education | 128 | 31.1 |
|  | College diploma and<br>above | 87 | 21.2 |
| Current occupation | Farmer | 6 | 1.5 |
|  | House wife | 134 | 32.6 |
|  | Private employee | 140 | 34.1 |
|  | Government<br>employee | 37 | 9.0 |
|  | Unemployed | 75 | 18.2 |
|  | Others | 19 | 4.6 |
| Current marital status | Married | 240 | 58.4 |
|  | Divorced | 19 | 4.6 |
|  | Separated | 17 | 4.1 |
|  | Widowed | 8 | 1.9 |
|  | Single | 127 | 30.9 |
| Family size | 5 & below | 333 | 81.0 |
|  | Above 5 | 78 | 19.0 |
| Household head | Male | 307 | 74.7 |

|  |  |  |
| --- | --- | --- |
| Female | 104 | 25.3 |

### Practice of respondents regarding iodized salt

Out of the total respondents, a quarter (24.8%), had poor practice towards iodized salt use. About two third (63.5%), of the participants reported that they were walking less than 30 minutes to get iodized salt, and half (52.1%) of the respondents reported that they were buying unpacked salt. Most (71.5%) of the respondents used a container with a cover to store salt at home, while almost three fourth (72.0%) of respondents were adding salt late at the end of cooking (Table 2).

**Table 2:**
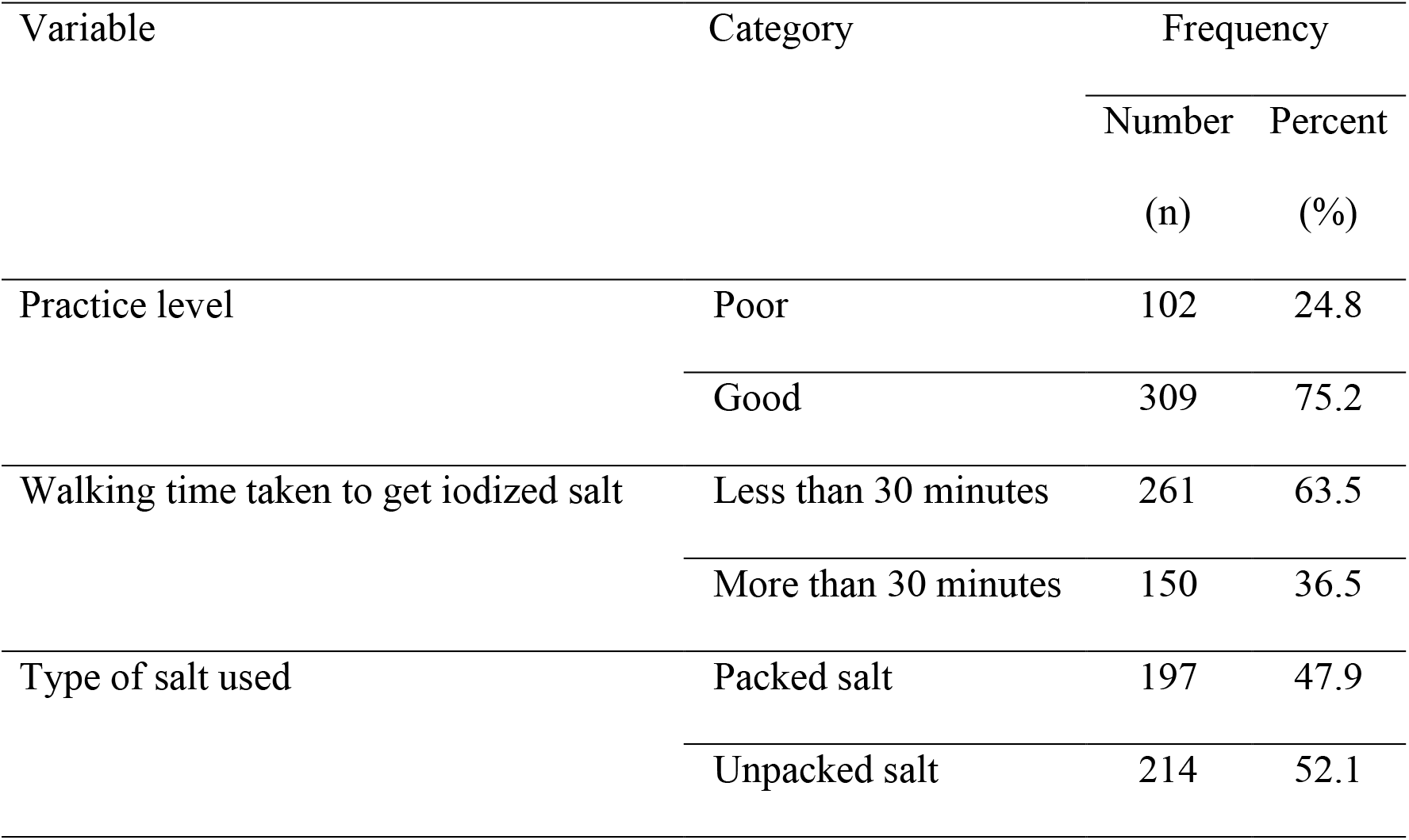
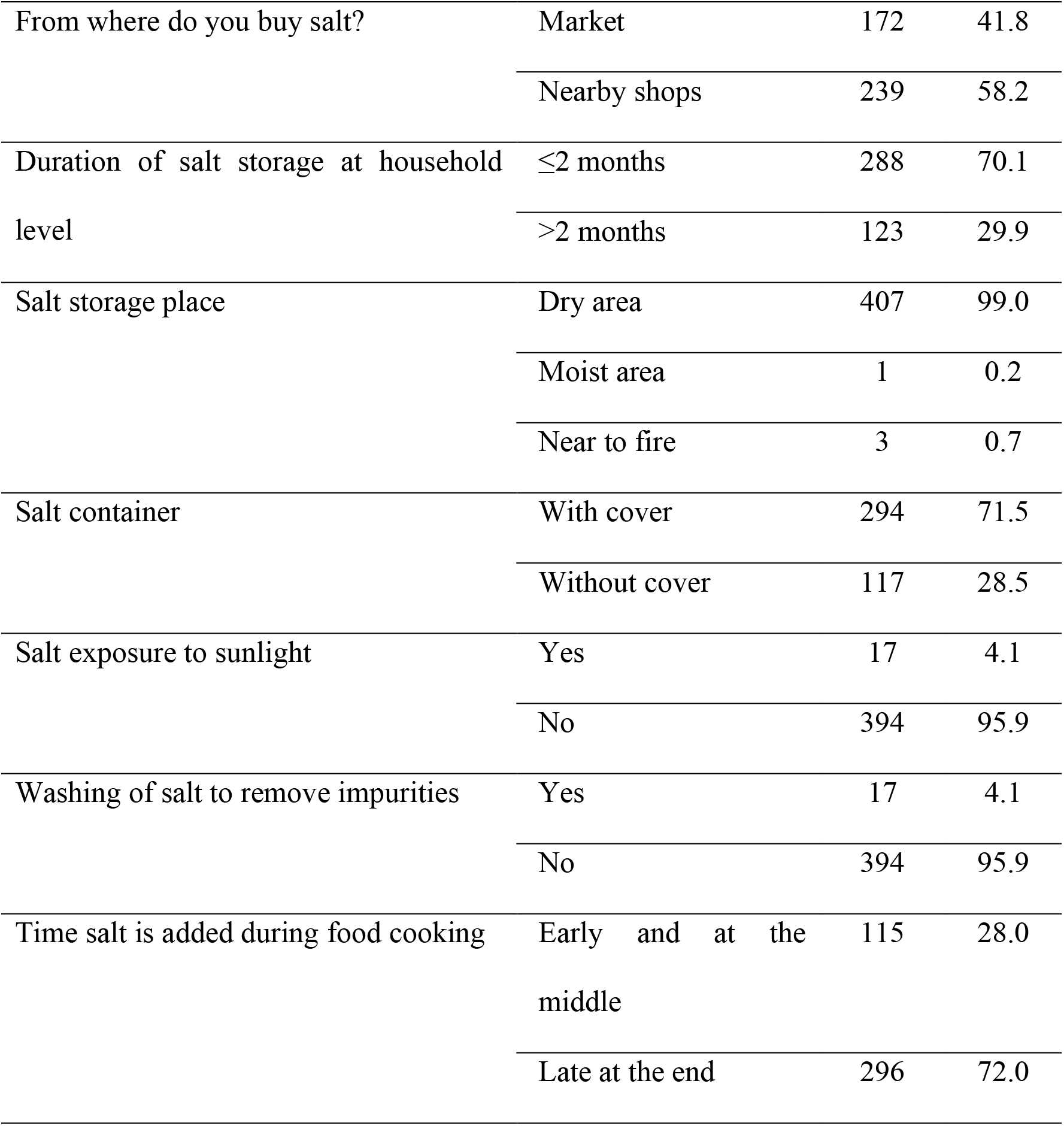
Practice of respondents regarding iodized salt in Gambela district (N=411).

### Availability of adequately iodized salt at household level

This study found that the availability of adequately iodized salt (15 PPM or more iodine) at household in Gambela district was 52.8% (95% confidence interval (CI): 0.47, 0.57). One hundred forty (34.06%) of respondents had inadequate iodine (less than 15 PPM) in their salt at household, whereas, 54 (13.14%) of them did not have any iodine (0 PPM) in their salt.

### Factors associated with availability of adequately iodized salt

From the bivariate logistic regression analysis, educational status of respondent, income, knowledge level, attitude level, walking time taken to get iodized salt, type of salt used, duration of salt storage, and salt container closure were associated (p-value < 0.25) with household availability of adequately iodized salt. However, during multivariate logistic regression analysis, only the following five variables were found to be positively associated with household’s availability of adequately iodized salt: educational status of respondent, attitude level, walking time taken to get iodized salt, type of salt used and salt container closure. Those participants with college diploma and above educational status were 4.27 times more likely to have adequately iodized salt contrary to those participants with no formal education (adjusted odds ratio (AOR)=4.27, 95% CI: 1.47, 12.44). Higher odds of availability of adequately iodized salt were observed among households with favorable attitude towards iodized salt use compared to those households with unfavorable attitude (AOR=5.69, 95% CI: 2.83, 11.42). Participants who walk less than 30 minutes to get iodized salt were 4.69 times more likely to have adequately iodized salt than their counterparts who walk more than 30 minutes (AOR=4.69, 95% CI: 2.56, 8.60). The odds of having adequately iodized salt were much higher among respondents who used packed salt compared to those used unpacked salt (AOR=2.75, 95% CI: 1.54, 4.93). Households with covered salt container were 3.93 times more likely to have adequately iodized salt contrary to those households without covered salt container (AOR=3.93, 95% CI: 2.21, 6.98) (Table 3).

**Table 3:**
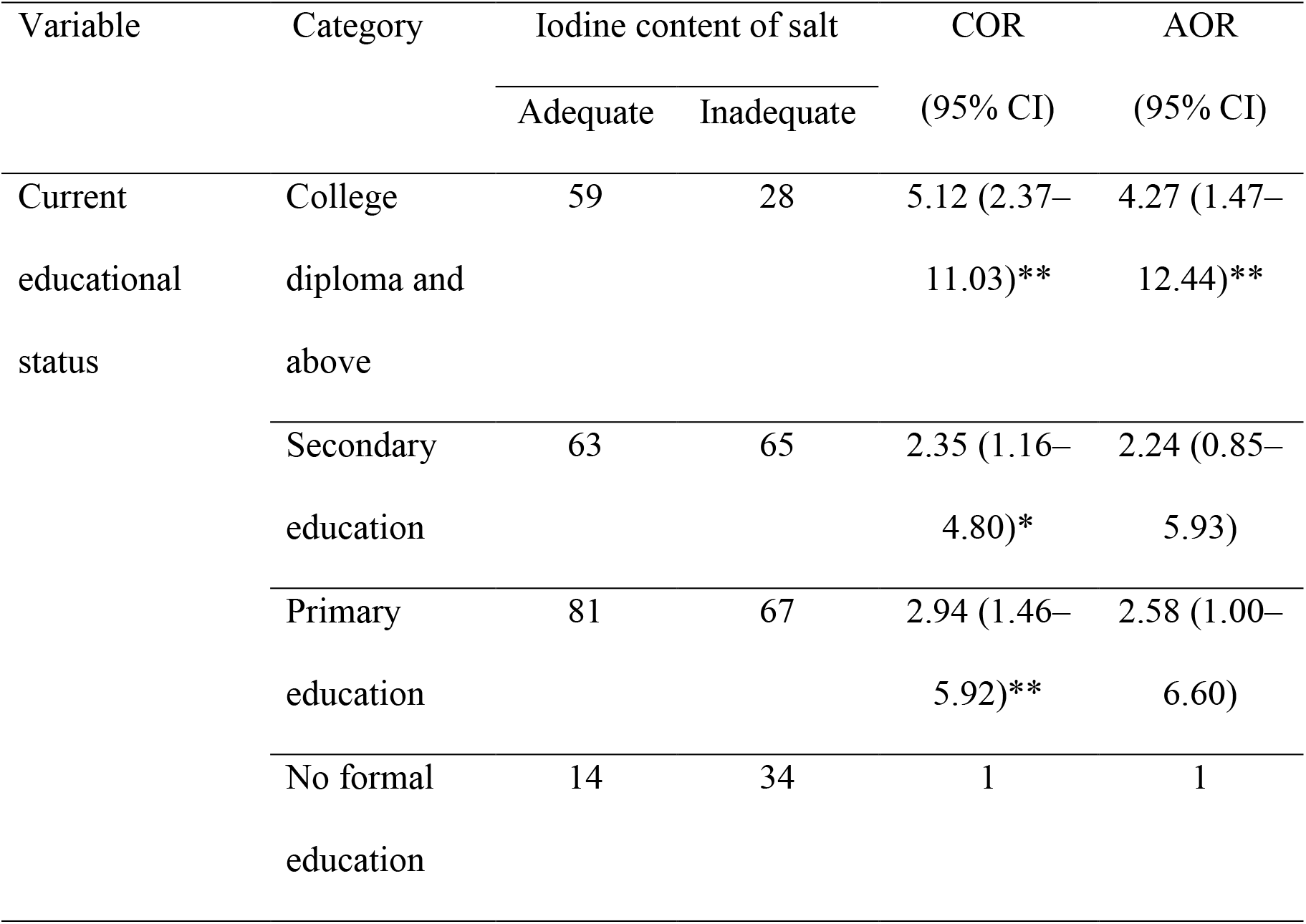
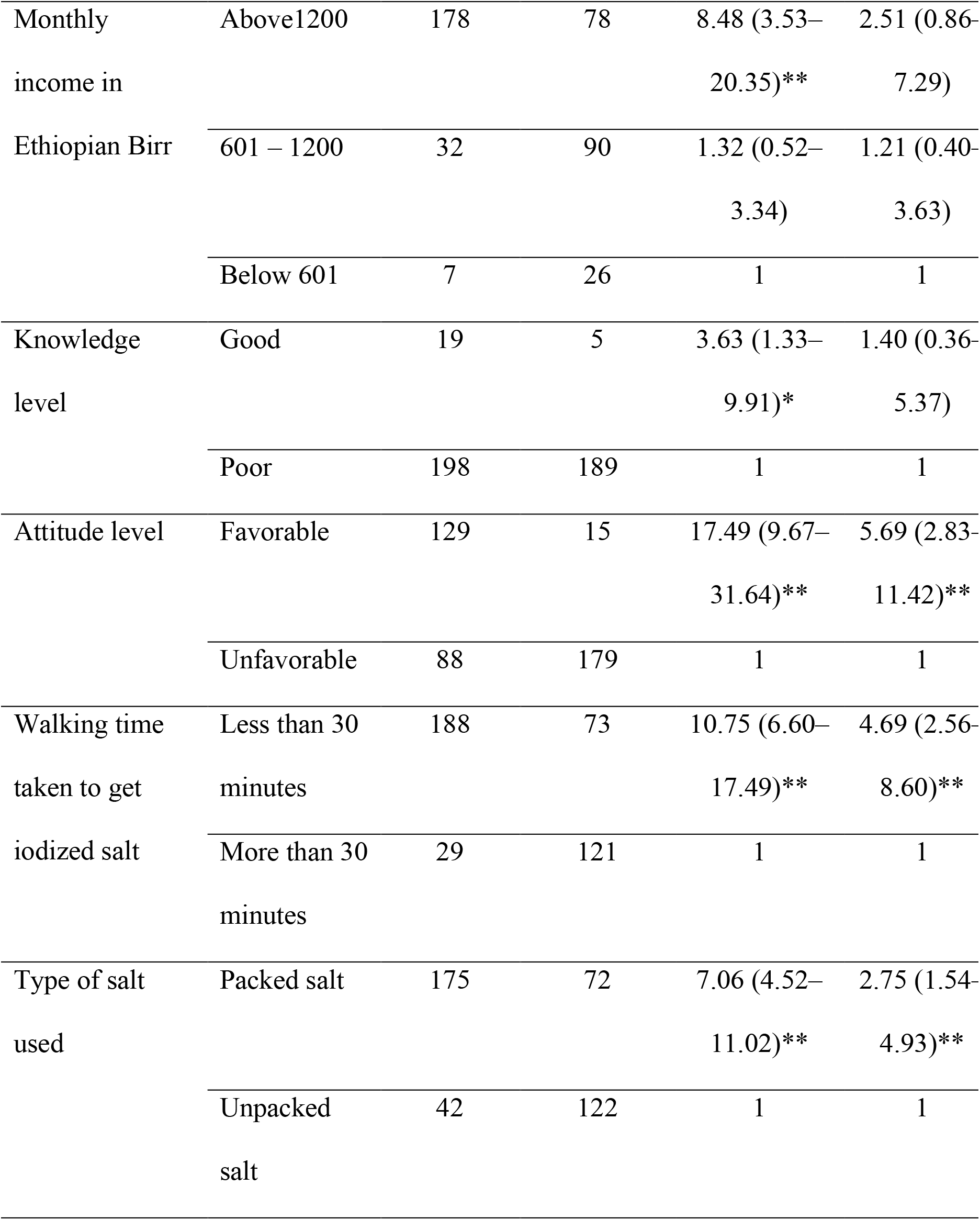
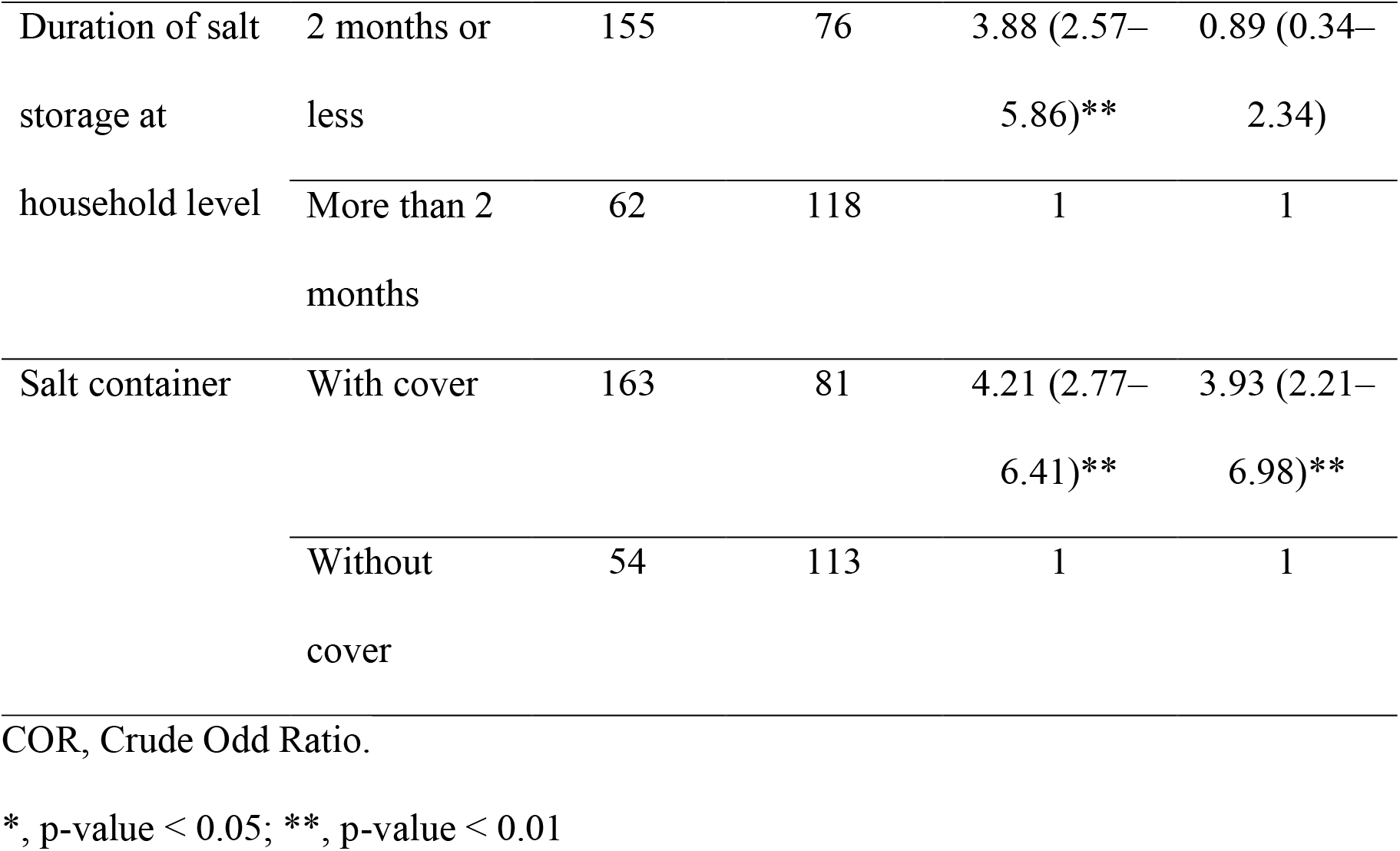
Factors associated with availability of adequately iodized salt in Gambela district (N=411).

## Discussion

USI was chosen as the best strategy for the prevention of iodine deficiency disorders since mostly people consume salt daily.^1^ Thereby, it was crucial to identify the level of iodine in a household’s salt. This study found that the availability of adequately iodized salt at households in Gambela district was 52.8% (95% CI: 0.47, 0.57). This finding is lower than the WHO USI target of 90% coverage.^7^ Nonetheless, the finding was higher compared to studies conducted in Dire Dawa town, Eastern Ethiopia (7.5%), in Robe town, South Eastern Ethiopia (32.7%), and in Kenya (26.2%) (23,27,31). This discrepancy is probably due to improved effort of local government to ensure market availability of iodized salt, and might also be due to more households in this study area store salt at a dry place than the latter studies.

The higher odds of availability of adequately iodized salt were observed among participants with college diploma and above educational status compared to those participants with no formal education. This association was also observed in studies done in Pakistan, in, Ghana and in Iraq.^17,18,32^ Furthermore, the finding was in line with studies in Ethiopia which indicated that having no formal education is risk factor for unavailability of adequately iodized salt.^20,33,34^

Those participants favorable attitude towards iodized salt use were more than five times more likely to have adequately iodized salt at household contrary to those with unfavorable attitude. The current finding is in agreement with a study carried out in Bangladesh, which showed that use of adequate iodized salt was more than double among participants with a positive attitude towards iodized salt.^35^

This study revealed that participants who walk less than 30 minutes to get iodized salt were more than four times more likely to have adequately iodized salt than those participants who walk more than 30 minutes. Another study conducted in Dabat district of Ethiopia also showed similar direction of association to the current study in that.^12^ A study in North Ethiopia indicated that iodized salt which is easily available or accessible to a household is positively associated with household’s iodized salt utilization.^36^

Various studies conducted in Ethiopia and Iraq revealed that using packed salt increases the chance of having adequately iodized salt at household.^23,32,37^ This study also showed that the odds of having adequately iodized salt were much higher among respondents who used packed salt compared to those used unpacked salt. The association could be due to packing iodized salt minimizes exposure to sunlight which eventually reduces iodine loss in the iodized salt.^29^

Households that were storing salt in a covered container were about four times more likely to have adequately iodized salt compared to those households that do not store salt in a covered container. Similarly, a study in Kenya found that households with covered salt container were more likely to have adequately iodized salt contrary to those households without covered salt container.^31^ This finding further reinforced by other studies conducted in Ethiopia.^22,25,37^ This association might be due to volatile nature of iodine when exposed to humidity, moisture and light while the iodized salt is stored in uncovered container that could lower the iodine level.

### Conclusion and Recommendation

The prevalence of availability of adequately iodized salt among households in Gambela district was much lower than the WHO recommendation of 90% of households, which should alert the public authorities to monitor regularly the quality of salt at household level in the study area. Educational status of respondent, attitude level, walking time taken to get iodized salt, type of salt used and salt container closure were found to be factors associated with the availability of adequately iodized salt among households in Gambela district. Therefore, in general, it is crucial to educate the community about the importance of iodized salt use and consequences of iodine deficiency, and efforts should be exerted so that they have favorable attitude towards iodized salt use. Furthermore, informing the public about proper use of iodized salt such as storing iodized salt in closed containers and using packed salt is indispensable. Concerned bodies should make sure that iodized salt is accessible to households in their vicinity, with in thirty minutes of walking.

## Data Availability

All data produced in the present study are available upon reasonable request to the corresponding author.

## Authors’ contributions

G.A.T. and E.N.G. conceived and designed the study. G.A.T., E.N.G., A.Y.U. and F.T.C. developed data collection tool, supervised data collection, and drafted the manuscript. G.A.T. and E.N.G. involved in data analysis and interpretation. G.A.T., E.N.G., A.Y.U. and F.T.C. drafted the manuscript, provided critical review and approved the final manuscript. All authors discussed the results and contributed to the final manuscript.

## Abbreviations

AOR: Adjusted Odds Ratio
CI: Confidence Interval
IDD: Iodine Deficiency Disorder
PPM: Parts Per Million
WHO: World Health Organization
USI: Universal Salt Iodization

